# Booster dose of BNT162b2 in a CoronaVac primary vaccination protocol improves neutralization of SARS-CoV-2 Omicron variant

**DOI:** 10.1101/2022.03.24.22272904

**Authors:** Guilherme R. F. Campos, Nathalie Bonatti Franco Almeida, Priscilla Soares Filgueiras, Camila Amormino Corsini, Sarah Vieira Contin Gomes, Daniel Alvim Pena de Miranda, Jéssica Vieira de Assis, Thaís Bárbara de Souza Silva, Pedro Augusto Alves, Gabriel da Rocha Fernandes, Jaquelline Germano de Oliveira, Paula Rahal, Rafaella Fortini Grenfelle Queiroz, Maurício L. Nogueira

## Abstract

The emergence of the new SARS-CoV-2 Omicron variant, which is known to accumulate a huge number of mutations when compared to other variants, brought to light the concern about vaccine escape, especially from the neutralization by antibodies induced by vaccination. In this scenario, we evaluated the impact on antibody neutralization induction, against Omicron variant, by a booster dose of BNT162b2 mRNA vaccine after the CoronaVac primary vaccination scheme. The percentage of seroconverted individuals 30 and 60 days after CoronaVac scheme was 17% and 10%, respectively. After booster dose administration, the seroconvertion rate increased to 76.6%. The neutralization mean titer against Omicron in the CoronaVac protocol decreased over time, but after the booster dose, the mean titer increased 43.1 times, indicating a positive impact of this vaccine combination in the serological immune response.

---

The vaccines against COVID-19 presented an important impact reducing the number of cases, hospitalization and deaths worldwide. However, in some countries the access to vaccination is far from the ideal, where the spread of SARS-CoV-2 continues at high rates favoring the emergence of variants. Indeed, since the beginning of the pandemics, the world has faced a continuous occurrence of variants, which is a natural process for a highly transmissible virus like SARS-CoV-2. Among these variants, the increase of transmission rate, augmented severity of cases and possibility to escape from immune response, stimulated by vaccination or previous infection, are the main concerns surrounding the called variants of concern (VOCs) [1, 2]. The omicron variant (B.1.1.529) has called attention due to its high number of mutations across the whole genome when compared to other VOCs. The omicron variant accumulates at least 47 mutations in its whole genome (the highest number among variants), of which more than half are present in the Spike protein, the main target of the serological immune response induced by the majority of COVID-19 vaccines [3-5].

In a global panorama, Brazil stands out as one of the epicenters for viral dissemination, presenting high numbers of cases and deaths. Along with USA and India, Brazil ranks in the top three of the most impacted countries, with more than 25 million cases and 625,000 deaths until February 7, 2022 [6]. On the other hand, historically, Brazil has a solid immunization program, which leaded to a huge SARS-CoV-2 vaccination adherence when compared to other countries, reaching more than 352 million doses administrated in the population, and almost 152 million people fully vaccinated. Among the distributed vaccines in Brazil, CoronaVac, the most used vaccine worldwide, was approved by our regulatory agency in January of 2021. This vaccine uses an inactivated virus technology and was the most administrated vaccine in Brazil until the middle of 2021, inoculated especially in elderly and healthcare workers individuals [7, 8]. The scheme consisted in a two doses protocol, with 2-4 weeks between doses. It is estimated that, until January of 2022, around 85 million doses of CoronaVac were administrated in the Brazilian population. With the introduction of some VOCs in the country, especially Delta (B.1.617.2) and Omicron variants, the Brazilian government adopted the distribution of a booster dose of the mRNA BNT162b2 vaccine (BioNTech/Pfizer) to enhance immune protection against COVID-19 [9].

In this context, our study evaluated the impact of the booster dose with BNT162b2 vaccine in a Brazilian cohort, under the CoronaVac two doses scheme, upon the stimulation of neutralizing antibodies against Omicron variant (HIAE – W.A). The participants were inoculated with two 0.5 ml shots of CoronaVac (600 SU per dose) in the original protocol, receiving a 0.3 ml booster of BNT162b2 (30 μg of spike mRNA per dose). A total of 90 individuals, randomly selected, were included in the study, divided equally into 3 different groups: The D30 group, where serum samples were collected 30 days after CoronaVac second dose; the D60 group, composed by samples collected 60 days after second dose; and D270 group, a time-point where samples were collected 270 days after second dose, comprehending also 30 days after mRNA booster dose.

Using a viral microneutralization assay (VNT_50_) with SARS-CoV-2 omicron variant live virus, and analyzing the percentage of seroconvertion, our results showed that after 30 days of second dose (D30), CoronaVac immunization generated detectable neutralizing antibodies against Omicron in only 17% of the included individuals. When comparing to the group collected 60 days after second dose (D60), the number of samples presenting neutralizing antibodies dropped to 10%, showing that, as expected, the serological immune response level, that was already low for the omicron variant, continued to decrease. However, after administration of BNT162b2 booster dose (D270), the number of samples presenting neutralizing antibodies against omicron significantly increased, reaching almost 77% of the included participants (Figure 1). The evaluation of VNT_50_ mean titers, for each time-point, shows that for D270, the mean titer was increased 27.5 times when compared with the mean titer observed for D30. In comparison with D60, this difference increases 43.1 times. These data show that the booster dose with BNT162b2, after CoronaVac vaccination, has improved the production of neutralizing antibodies against the Omicron variant. The neutralizing improvement here described could be explained by the fact that beta-coronaviruses, including SARS-CoV-2 variants, share a conserved site into Spike protein. Since BNT162b2 vaccine uses Spike mRNA from the original Wuhan SARS-CoV-2 isolate, this conserved region could allow the recognition of Omicron variant by the induced antibodies, independently on the number of accumulated mutations [10].

**Figure 1.**
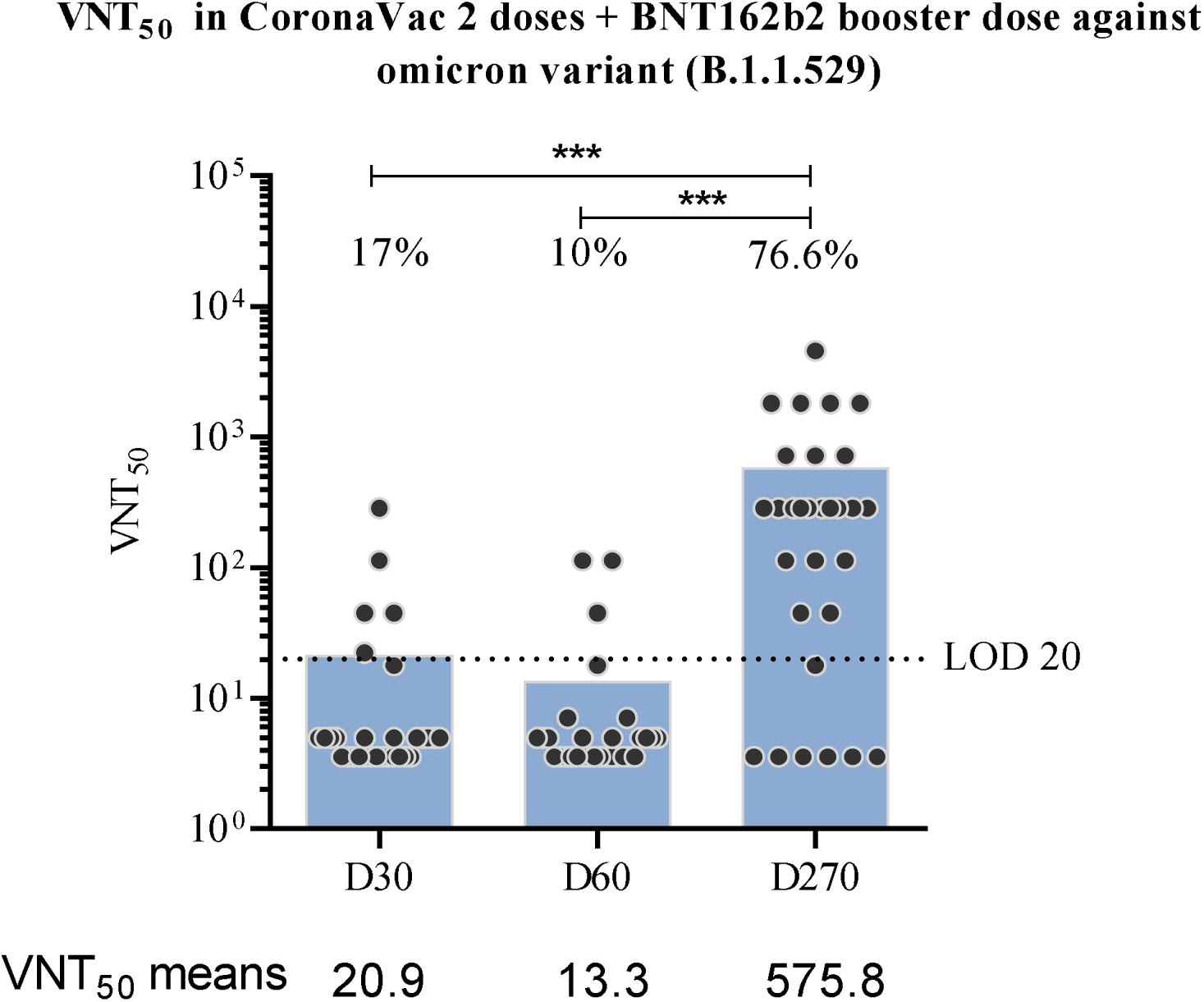
Viral microneutralization assay against SARS-CoV-2 omicron variant: 30 days after CoronaVac second dose (D30); 60 days after CoronaVac second dose (D60); 270 days after CoronaVac second dose and 30 days after BNT162b2 booster (D270). The dark grey circles represent the included participants, and the 50% neutralizing titer (VNT_50_) for each sample was assayed using the Omicron variant live virus (HIAE – W.A) (See the Supplementary Appendix for methods description). The blue bars represent the VNT_50_ mean values. The dashed line represents the lower limit of detection (LOD), determined in this assay as 20. The percentage of seroconverted samples, for each group, is indicated in the graph. The statistical analysis was obtained by one-way ANOVA and Tukey’s multiple comparisons test, where the three asterisks represent the P value < 0.0001.

It is worth to notice that our data corroborates with some findings recently published by GeurtsvanKessel and colleagues (2022) analyzing the BNT162b2 booster dose effect in other vaccination schemes, both in the serological and cellular immune response [11]. In the study, the authors evaluated four different immunization schemes, two using adenovirus-vector technologies and two using the mRNA platform (including BNT162b2 original scheme). They observed that neutralizing antibodies against Omicron variant were significantly lower or absent, depending on the used vaccine, when compared to VOCs like Beta and Delta. However, just as showed by our data, the booster dose with BNT162b2 restored neutralization titers against omicron for all the tested schemes. It is interesting to highlight that the cellular response against Omicron, mediated by T-cells, was maintained in all the tested vaccination protocols, showing that the highest impact of this variant in the immune response is mainly in the antibody neutralization ability [11].

From the clinical point of view, a recent article showed, in a Brazilian cohort, that administration of BNT162b2 after CoronaVac increased protection against infection when comparing to CoronaVac original protocol. The CoronaVac two doses scheme presented effectiveness of 34.7% against new infections when compared to unvaccinated individuals, and this protection showed to decrease over time. However, the administration of BNT162b2 booster was responsible to increased protection against infection to 82.6%. The same pattern was observed for COVID-19 disease progression, showing that the booster dose reduces the chances of severe outcomes, especially in risk groups [12]. Another study, also from the clinical area, but analyzing this booster after the BNT162b2 original vaccination scheme, showed a reduction of 90% in the mortality rate [13]. Once our results describe the protection improvement caused by BNT162b2 booster from the perspective of the serological response, all of these studies complement our observations, showing the impact of this immunization approach on distinct branches of the immune system.

As limitations of our study, the antibody neutralization analysis here performed was focused only in the Omicron variant. Also, it is well known that serological response is not the only immunological aspect that is stimulated by the vaccination, and that cellular immunity, in association with clinical data, is required to successfully determine the effectiveness of a vaccine. Nevertheless, the data here presented are strengthened by the study of GeurtsvanKessel and collaborators [11], emphasizing the need for constant monitoring of neutralizing antibody capacity, especially given the possibility of novel variants emerging in the future.

Thus, our data shows that, against Omicron variant, neutralization titers induced by primary vaccination with CoronaVac decreased with time and was 43.1 times lower when compared to the use of a booster dose with BNT162b2. Therefore, this neutralization enhancement after booster dose could reflect, clinically, in higher protection and lower risk of disease progression.

## Data Availability

All data produced in the present work are contained in the manuscript

