## Supplementary appendix for "Booster dose of BNT162b2 in a CoronaVac primary vaccination protocol improves neutralization of SARS-CoV-2 Omicron variant"

**Viral Microneutralization Assay**

In order to evaluate the antibody neutralization levels against SARS-CoV-2 omicron variant, in samples collected from a cohort vaccinated with the CoronaVac two doses protocol, followed by the BNT162b2 mRNA vaccine booster dose, a viral microneutralization (VNT) was performed. In a day before, 10^4^ Vero CCL-81 cells were distributed, per well, in clear flat bottom 96 well plates and incubated for 24h in a water-jacked incubator, at 37°C and 5% CO_2_, reaching 90-95% of confluence after incubation. Serum samples, collected from the cohort participants, were heat-inactivated for 30 min, at 56°C. To analyze the neutralization capacity of antibodies induced by vaccination, using microdilution 96 well plates, the inactivated samples were serially diluted in fresh DMEM media supplemented with 100 U/ml of penicillin, 100 µg/ml of streptomycin and 2% of fetal bovine serum (FBS). For each serum sample, eight dilutions were tested (1:20, 1:40, 1:80, 1:160, 1:320, 1:640, 1:1280 e 1:2480). The fresh diluted samples were incubated with the live SARS-CoV-2 Omicron variant (HIAE – W.A) in a constant concentration of 50 TCID50/ml, at 37°C, for 1h. After this period, the diluted samples, mixed with virus, were distributed in the Vero CCL-81 cell plates and maintained in the humidified incubator at 37°C and 5% CO_2_ for 72h. Thereafter, media containing serum and viruses was discarded, cells were fixed with 10% formaldehyde, for 20 min, and stained with crystal violet. The neutralization capacity was determined by the presence or absence of cytopathic effect across the dilutions, and for each sample, it was obtained the reciprocal dilution value where 50% of cytopathic effect was avoided (VNT_50_). All samples, at all dilutions, were tested in triplicate and VNT50 was calculated by the use of the Spearman-Karber algorithm [1, 2]. To validate the assay, a serum sample already known to neutralize Omicron and a sample collected before the pandemics were used as positive and negative control, respectively.

1.	Spearman, C., *The Method of “Right and Wrong Cases” (Constant Stimuli) without Gauss’s Formula.* Br J Psychol, 1908(2): p. 15.

2.	Kärber, G., *Beitrag zur kollektiven Behandlung pharmakologischer Reihenversuche.* Archiv f experiment Pathol u Pharmakol, 1931(162): p. 4.
